# Efficacy and Safety of Chinese Herbal Injection Thoracic Perfusion combined with Cisplatin chemotherapy for Lung Cancer Malignant Pleural Effusion: A Systematic Review and Meta-Analysis

**DOI:** 10.1101/2022.12.01.22283004

**Authors:** Zhong-Ning He, Ming-Wan Su, Jie-He, Guang-Hui Zhu, Yu-Wei Zhao, Shu-Lin He, Yue Li, Bo-Lun Shi, Jia-Qi Hu, Shun-Tai Chen, Bao-Jin Hua

## Abstract

**Purpose:** To evaluate the efficacy and safety of Chinese herbal injection thoracic perfusion combined with cisplatin chemotherapy for lung cancer patients with MPE.

**Method:** Eight databases were searched for articles published from inception to August 20, 2022 for randomized-controlled trials (RCTs) that are relevant. The included studies were reviewed by two investigators, with relevant data extracted independently. Primary outcome was identified as objective response rate (ORR), while secondary outcomes were identified as quality of life (QOL) and adverse reactions. Quality of the included trials was assessed through risk of bias assessment of the Cochrane risk-of-bias tool. The Revman5.3 and Stata17.0 software were used to calculate risk ratio (RR) at 95% confidence intervals (CI) for binary outcomes.

**Results:** 29 RCTs involving 1887 patients were included in this study. Compared with patients treated with cisplatin thoracic perfusion alone, those with Chinese herbal injection and cisplatin thoracic perfusion had better therapeutic effects (RR=1.44, 95% CI: 1.35∼1.53, P = 0.000), higher KPS score (RR = 1.47, 95%CI: 1.34∼1.61, P = 0.0000), lower digestive tract reaction(RR = 0.56, 95%CI: 0.48∼0.67, P = 0.000), bone marrow suppression (RR = 0.50, 95% CI: 0.43∼0.59, P =0.000) and chest pain reactions (RR = 0.65, 95%CI: 0.47∼0. 89, P = 0.007).

**Conclusion:** The systematic review indicated that Chinese herbal injection thoracic perfusion combined with cisplatin chemotherapy may improve therapeutic effect, quality of life, and reduce adverse reactions. More large-scale and higher quality RCTs are warranted to support our findings. Systematic Review Registration: PROSPERO, identifier CRD42022347345.

## 1 Introduction

Lung cancer is one of the most common malignant tumors in the world, with high morbidity and mortality^[1]^, Malignant Pleural Effusion (MPE) is a common complication in lung cancer patients. and the median survival time is only 5.5 months^[2]^. The occurrence of MPE is associated with poor quality of life (QOL) and shorter survival time in lung cancer patients^[3]^. Thus, effective treatment for MPE appears very important. The main purpose is to control the effusion from growth, relieve dyspnea, improve the quality of survival, to extend the survival. Currently, management options for MPE includes chest tube drainage and systematic chemotherapy to achieve symptomatic relief and controlling the primary malignancies. Thoracic perfusion of chemotherapeutic is an effective method for the treatment of MPE, and thoracic perfusion of cisplatin has good pharmacodynamics^[4, 5]^, high economic practicability^[6]^. However, the side effects of cisplatin, such as gastrointestinal reactions and hepatorenal toxicity, cannot be ignored, and will affect the quality of life of patients to a certain extent^[7]^. Therefore, current management approach remain palliative. Thus, we seek to find an effective combined treatment for MPE. In recent years, related studies have shown that anti-tumor Chinese herbal injection thoracic perfusion combined with cisplatin chemotherapy in the treatment of MPE have a positive effect in improving efficiency, improve the quality of life, reduce adverse reactions^[8-10]^. Although the clinical studies on Chinese herbal injection in the treatment of lung cancer MPE are increasing, the long-term efficacy has not been measured. Lack of high quality, neat standard clinical research, In order to obtain higher evidence data, this study using evidence-based medicine method, comprehensive collection published in Chinese and English literature at present, the systematic evaluation of Chinese herbal injection thoracic perfusion combined with cisplatin chemotherapy of lung cancer MPE, curative effect in order to provide a basis for evidence based medicine in the future clinical treatment.

## 2 METHODS

This study was conducted following the Preferred Reporting Items for Systematic Reviews and Meta-Analyses (PRISMA) guidelines^[11]^. The protocol has been registered in PROSPERO (registration number: CRD42022347345).

### 3 Search strategy

An independent review of citations was searched for articles published from inception to August 20, 2022, relevant publications evaluating the efficacy and safety of Chinese herbal injection for Lung cancer with MPE were searched in the Chinese National Knowledge Infrastructure (CNKI), Wanfang Database, the VIP Information Database, Chinese Biomedical Literature database (CBM), PubMed, EMBASE, and the Cochrane Library. The following Key words were used: “Lung Neoplasms OR Lung Cancer” AND “Pleural Effusion, Malignant OR Malignant Pleural Effusion” AND “Cisplatin” AND “Elemene injection OR Kushen injection OR Kanglaite injection OR Brucea Javanica oil emulsion OR Yuxingcao injection OR Aidi injection OR Xiao-ai-ping injection OR Huachansu injection OR Shenqifuzheng injection OR Kang-Ai injection OR Kushenhuangqi injection”. All retrievals were implemented using MeSH and free words (the detailed search strategy is available in Supplementary 1). The languages were restricted to Chinese and English.

### 4 Study selection and data extraction

Eligible studies were randomized controlled trials of lung cancer adults (≥18) with MPE involvingChinese herbal injection thoracic perfusion combined with cisplatin chemotherapy comparing to cisplatin thoracic perfusion alone. We defined intervened injections as Elemene injection, Kushen injection, Kanglaite injection, Brucea Javanica oil emulsion, Yuxingcao injection, Aidi injection, Xiao-ai-ping injection, Huachansu injection, Shenqifuzheng injection, Kang-Ai injection, Kushenhuangqi injection etc. Studies were excluded if patients were treated with other chemotherapy.

Two reviewers Ming-Wan Su and Jie-He independently selected studies based on the inclusion and exclusion criteria described above. Two reviewers independently extracted details for each trial: first author name and year of publication, patient characteristics; Intervention: Chinese herbal injection, Cisplatin ingredients, dose, treatment time; primary and secondary outcome measures. Missing information can be obtained by contacting the author.

### 5 Data synthesis and analysis

The primary outcome measure is objective remission rate (ORR) of MPE which was defined as CR + PR^[12]^. Quality of life (QOL) evaluated with Karnofsky score. Adverse events (digestive tract reactions, bone marrow suppression, chest pain) are second outcome measures. The data were processed and analyzed by Stata 17.0 (Stata Corp, College Station, SE). Random-effects models were performed to calculate pooled effects. Fixed-effect models were performed if statistical heterogeneity was absent (heterogeneity test, P ≥ 0.10), dichotomous data were presented as pooled Risk Ratio (RR) with 95% confidence intervals (95% CIs), Assessment of heterogeneity was performed using Cochran’s Q test and Higgins’s I^2^; I^2^ > 50% and a P value < 0.10 suggested significant heterogeneity^[13]^. Sensitivity analysis was performed by sequentially omitting each study to examine the robustness of the results. Potential publication bias was evaluated using Begg’s funnel plot and Egger’s test^[14]^.

### 6 Risk of Bias Assessment

Two reviewers Zhong-Ning He and Guang-Hui Zhu independently assessed the quality of the selected trials according to the RCT trials tool of the Cochrane Collaboration. Items were divided into three categories: low risk bias, unclear bias and high-risk bias. The following features are random sequence generation (selection bias), concealment of assignment (selection bias), blinding of participants, personnel (performance bias), outcome data were incomplete (attrition bias), selective reporting (reporting bias), and other bias. These assessments are plotted and evaluated using Review Manager 5.3.

## 7 Result

### 7.1. Search Result

Initially 281 literatures from 7 electronic databases were identified. After the removal of 98 duplications and 55 irrelevants after screening through the title and abstract, 43 articles were consider potentially appropriate for inclusion. Among which, 14 articles were excluded for the following reasons: not lung cancer patient (n=2), not thoracic perfusion (n=4), Chinese herbal injection intervention alone (n=6), Chinese herbal injection with other chemotherapy (n=2). Eventually, 29 single-centered RCTs met all inclusion criteria,^[8-10, 15-40]^ as shown in the flow diagram (Locate Figure 1)

**Figure 1.**
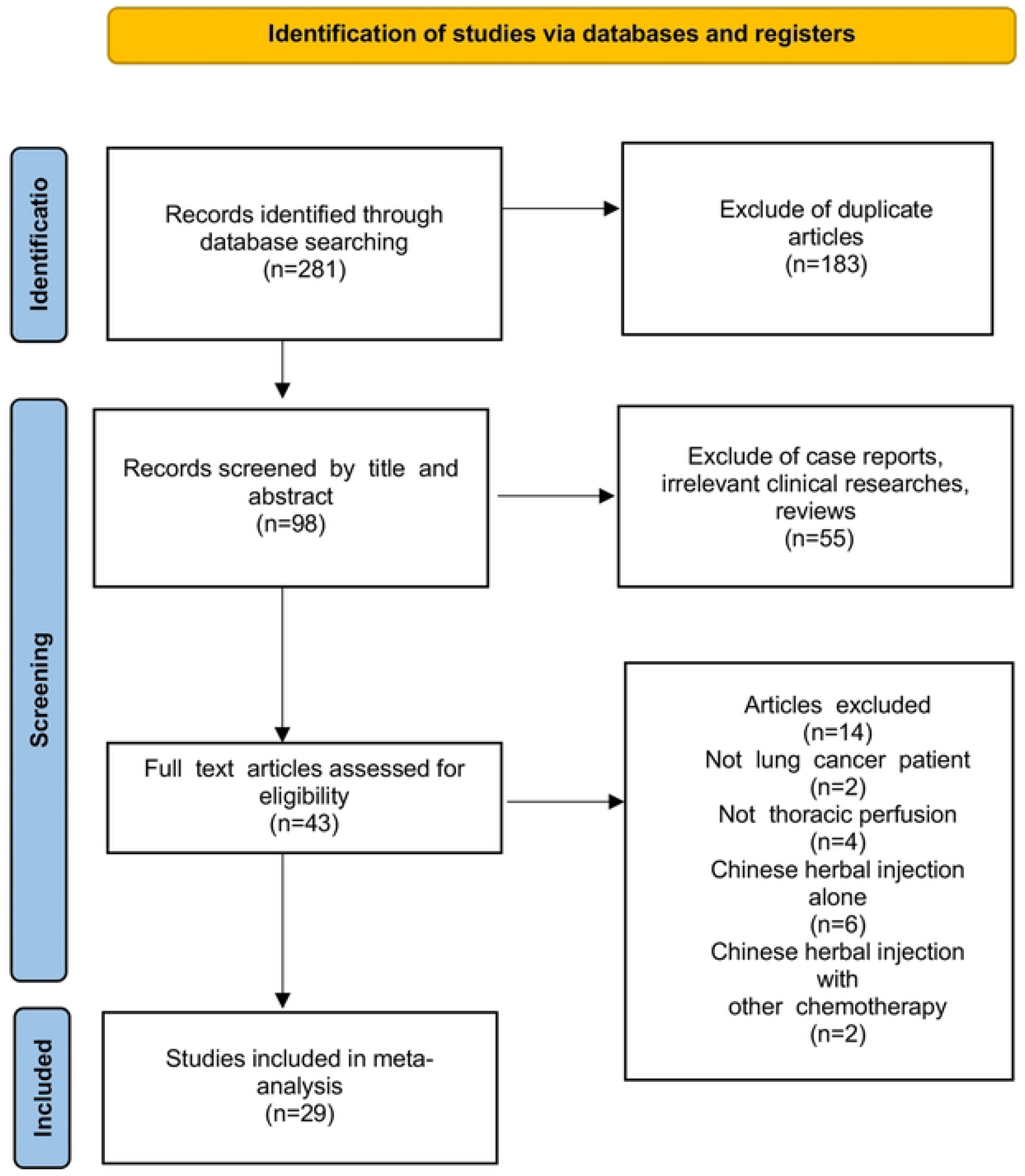
Flow diagram

### 7.2. Characteristics of the Included Studies

All 29 studies yielded a total of 1,887 patients. The baseline characteristics of each trial are presented in Table 1. The control group was treated with cisplatin thoracic perfusion, on the basis of the control group, the experimental group was infused with chinese herbal injection. 29 trials reported the ORR following WHO guidelines, KPS was reported in 16 studies, and adverse events were reported in 24 studies, Among the 29 citations, intervention of experimental group was Aidi injection in 12 studies^[10, 18, 21, 24, 26, 30-32, 35-38]^, Kushen injection in 10 studies^[8, 16, 17, 20, 22, 25, 27, 33, 34, 40]^, Brucea Javanica oil emulsion in 3 studies^[9, 29, 39]^, and other injections were not included because less than 3 studies. A total of 16 studies reported QOL improvement (the number of patients KPS score 10 points higher or stable after treatment)^[8, 9, 15, 16, 21, 22, 24-27, 29, 33, 34, 36, 37, 39]^. 24 studys that included 1,527 patients reported adverse reactions^[8, 9, 16-27, 29, 31-39]^. Bone marrow suppression was recorded in 23 studies^[8, 9, 16-21, 23-27, 29-32, 34-39, 41]^. 19 studys that included 1,353 patients reported chest pain^[9, 15, 16, 8-24, 26, 29, 31-33, 35, 37-39]^.

**Table1.**
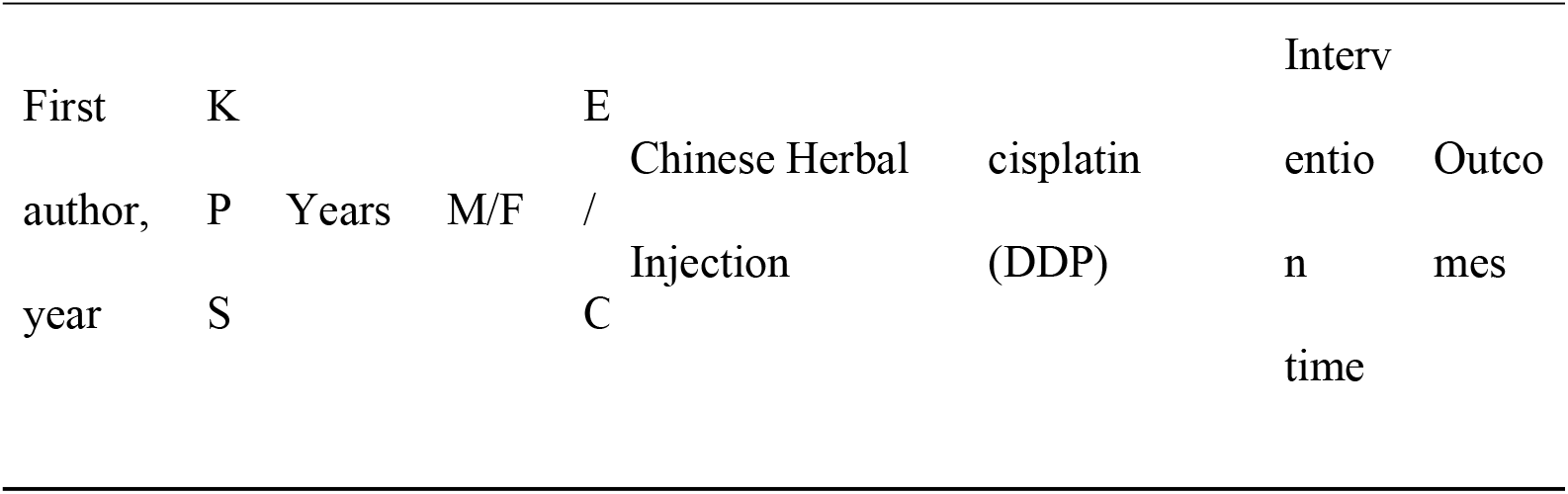
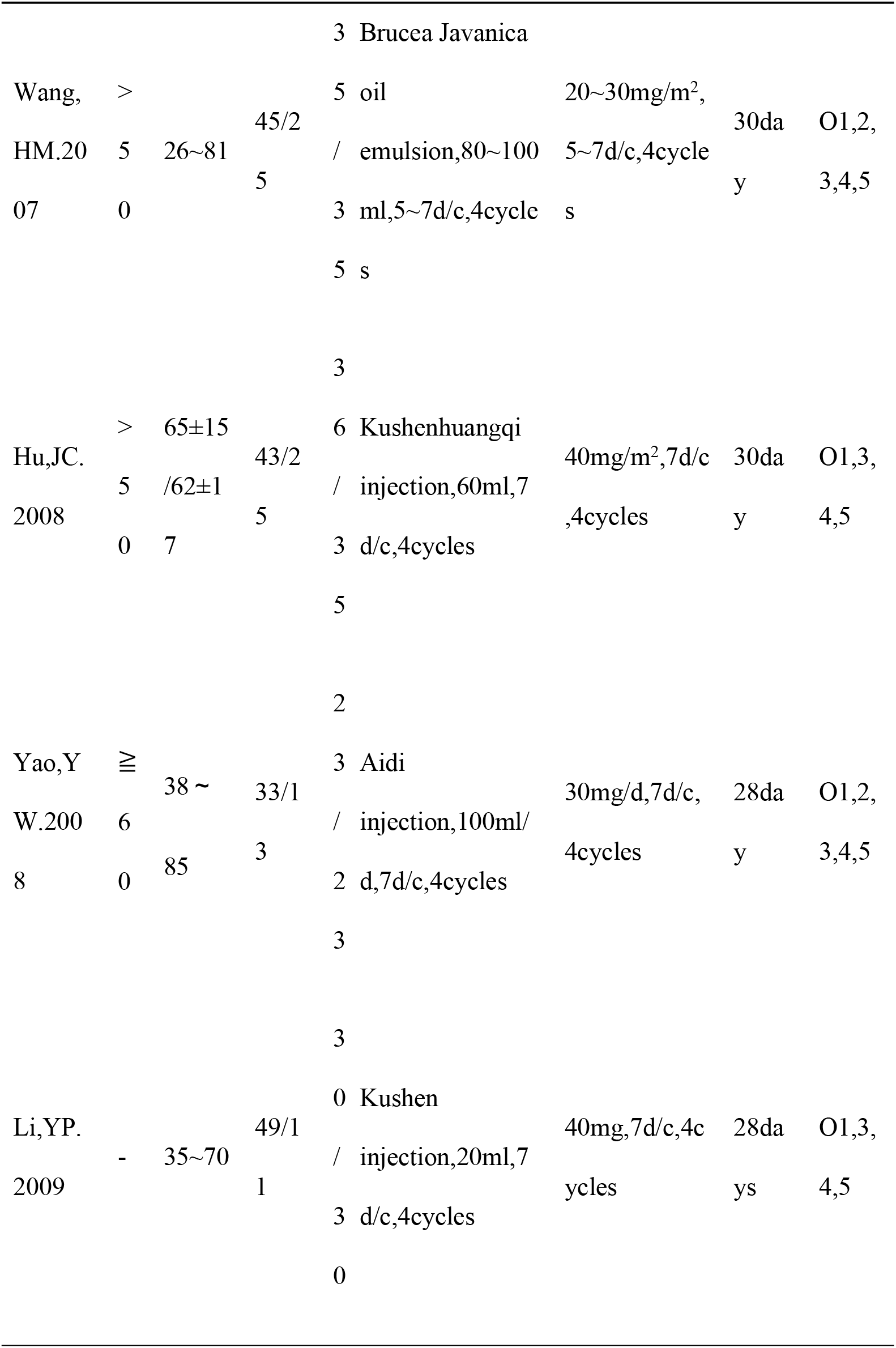
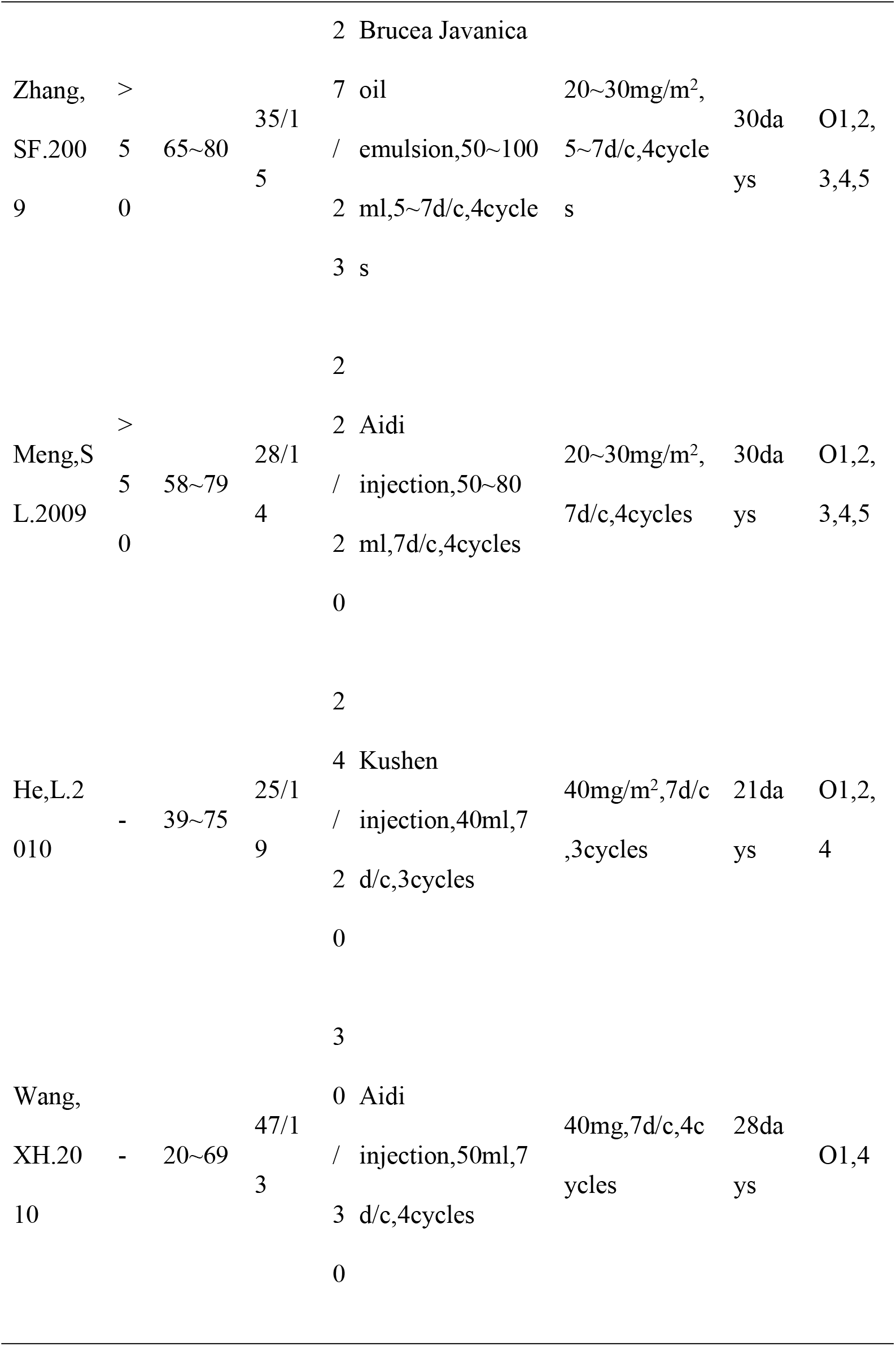
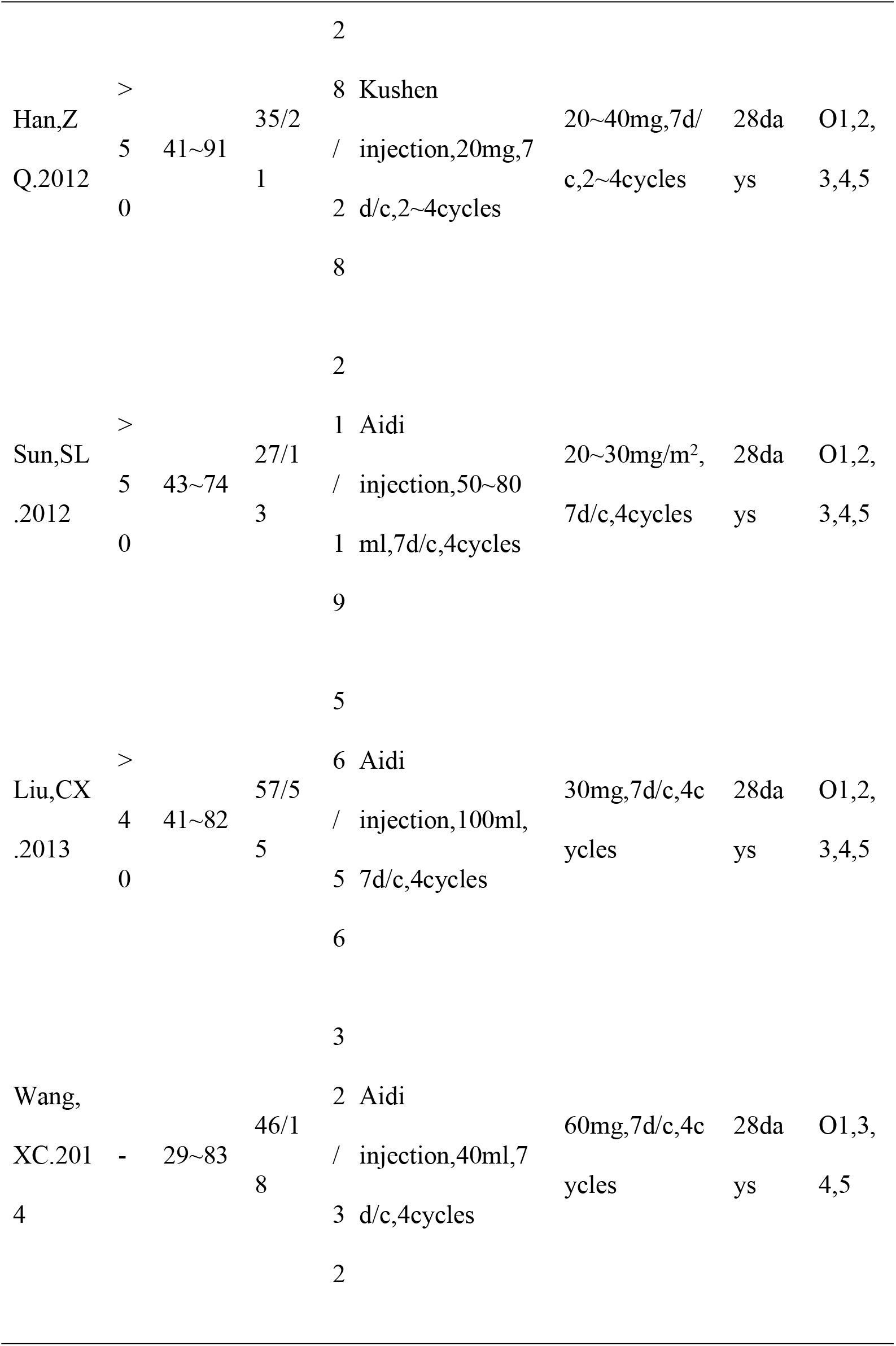
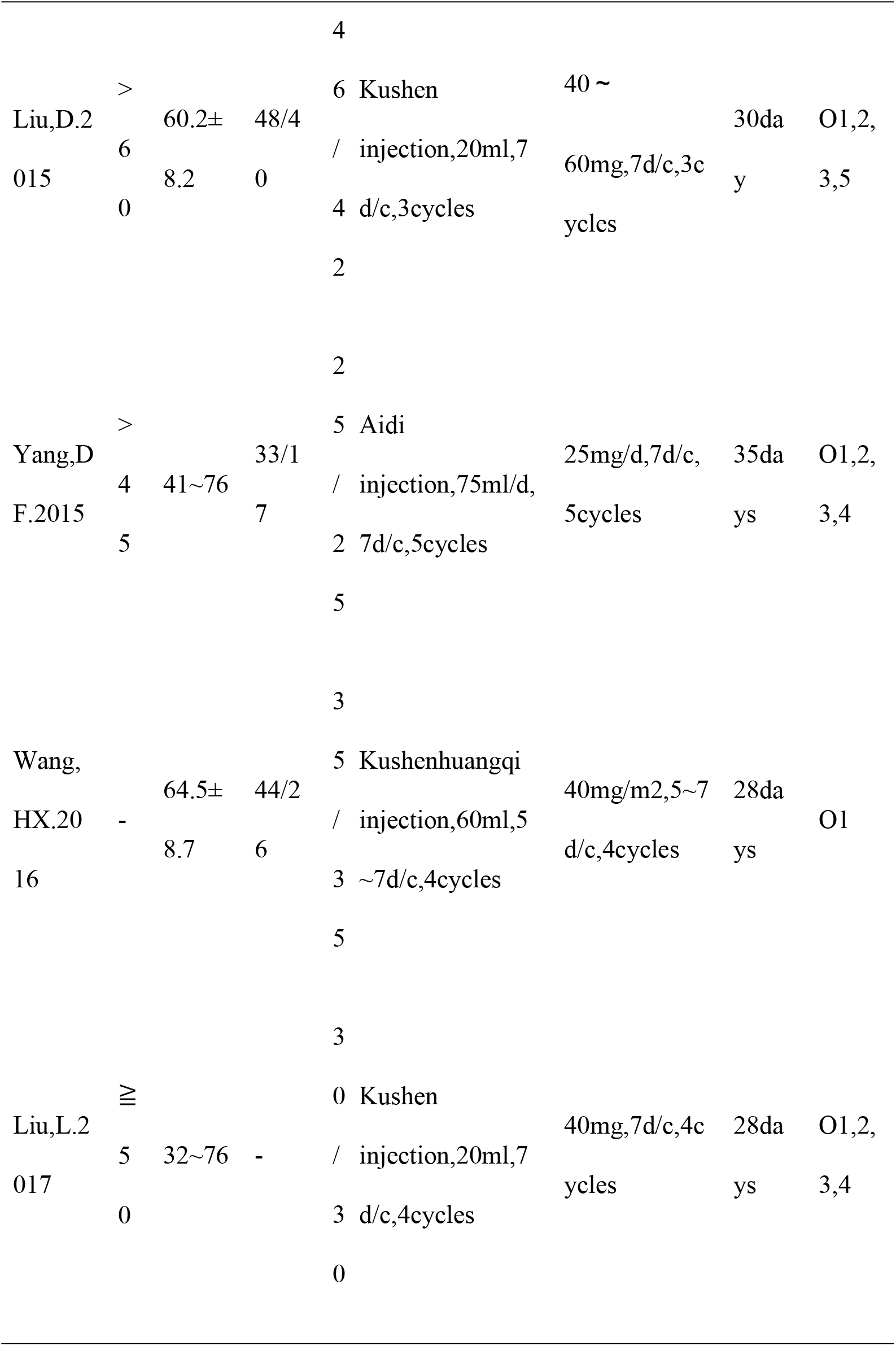
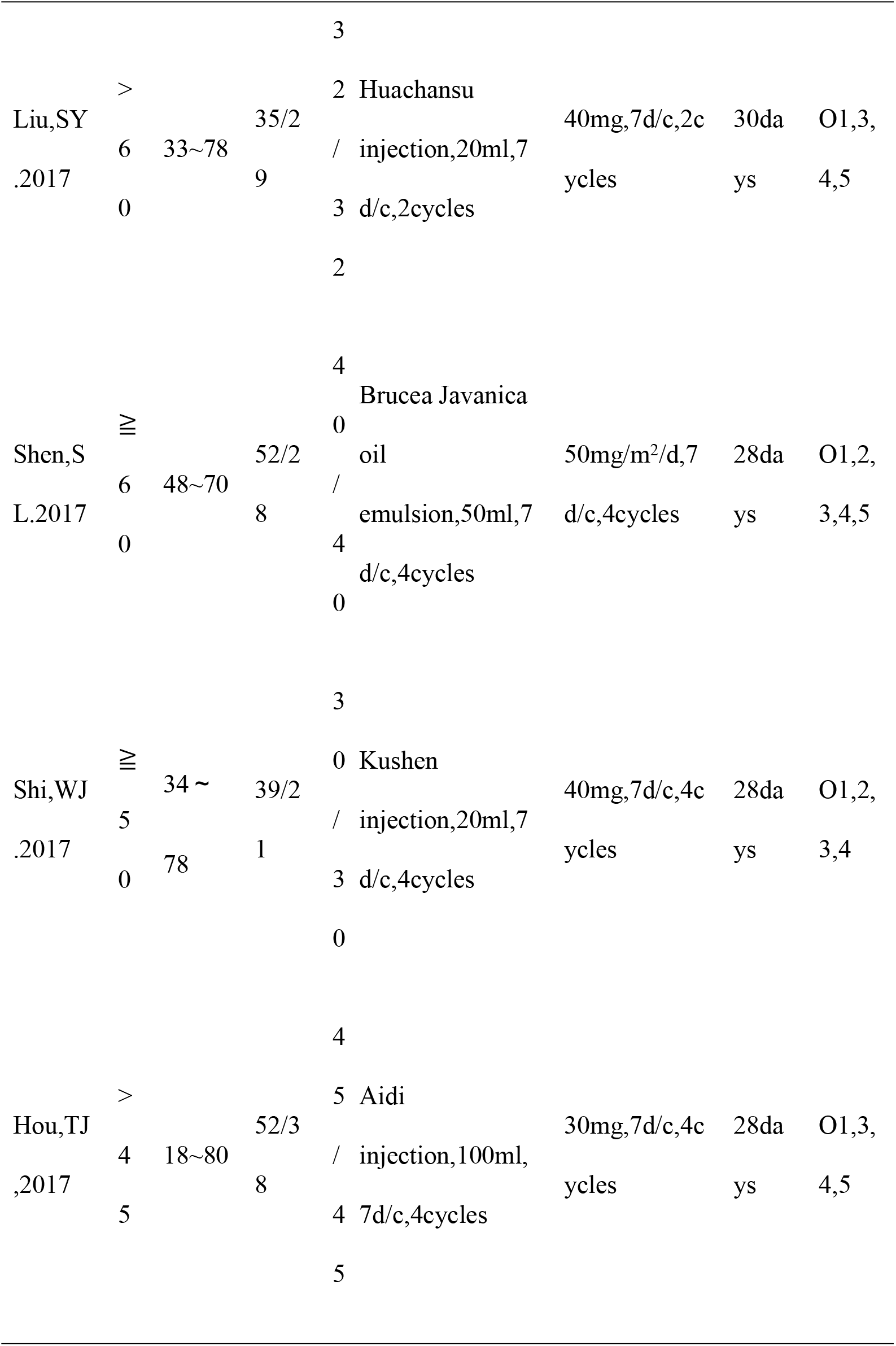
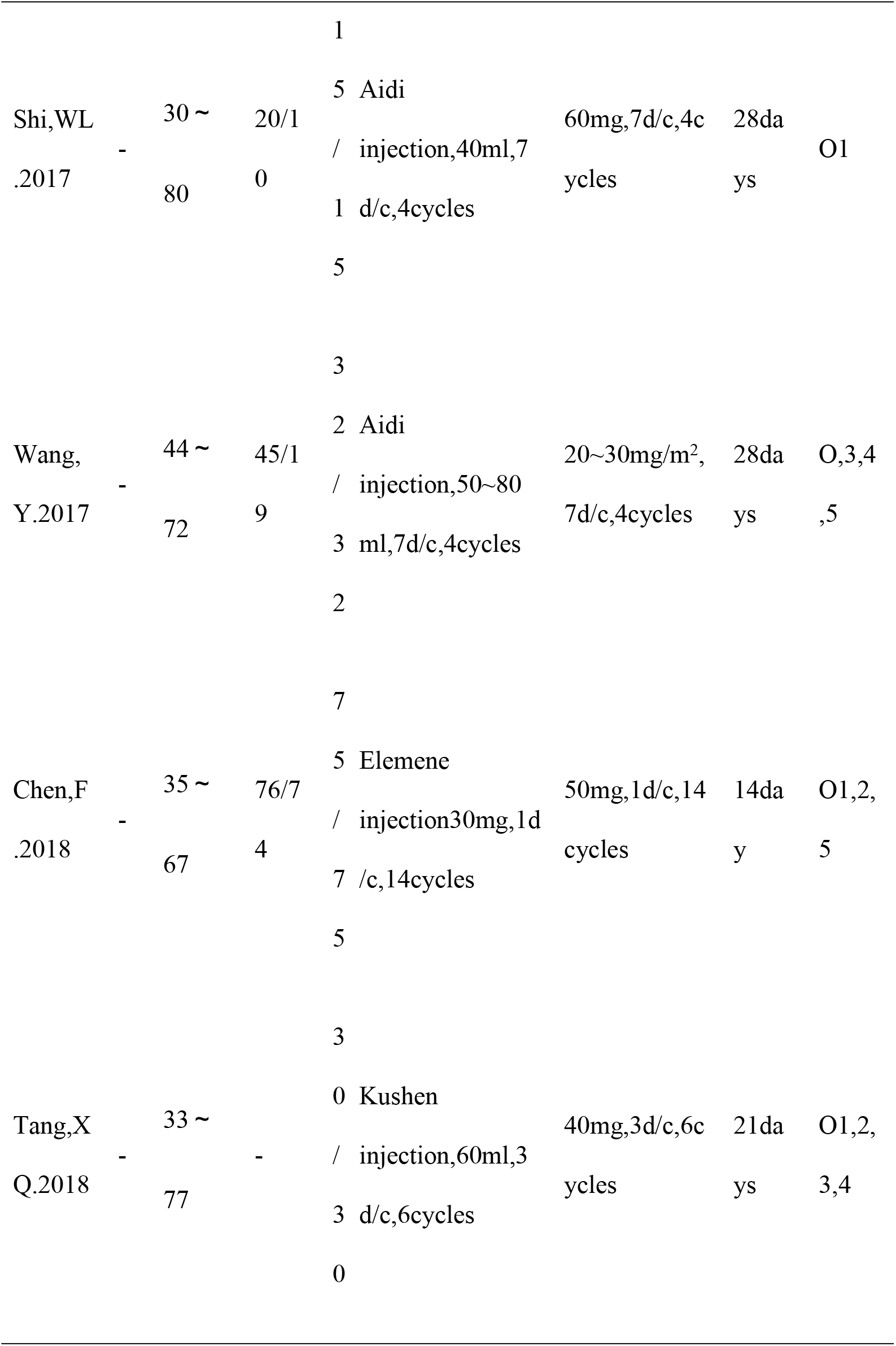
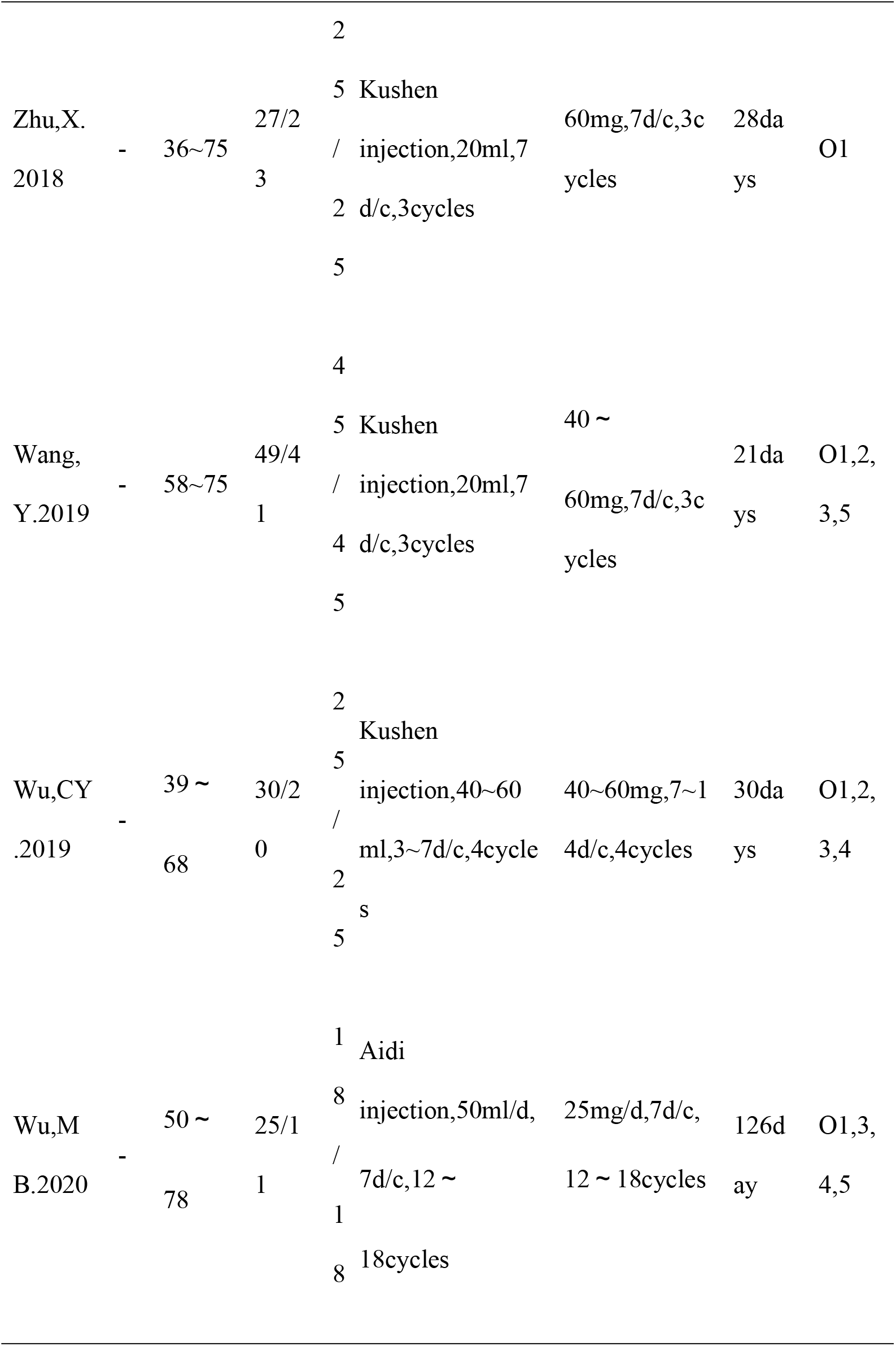
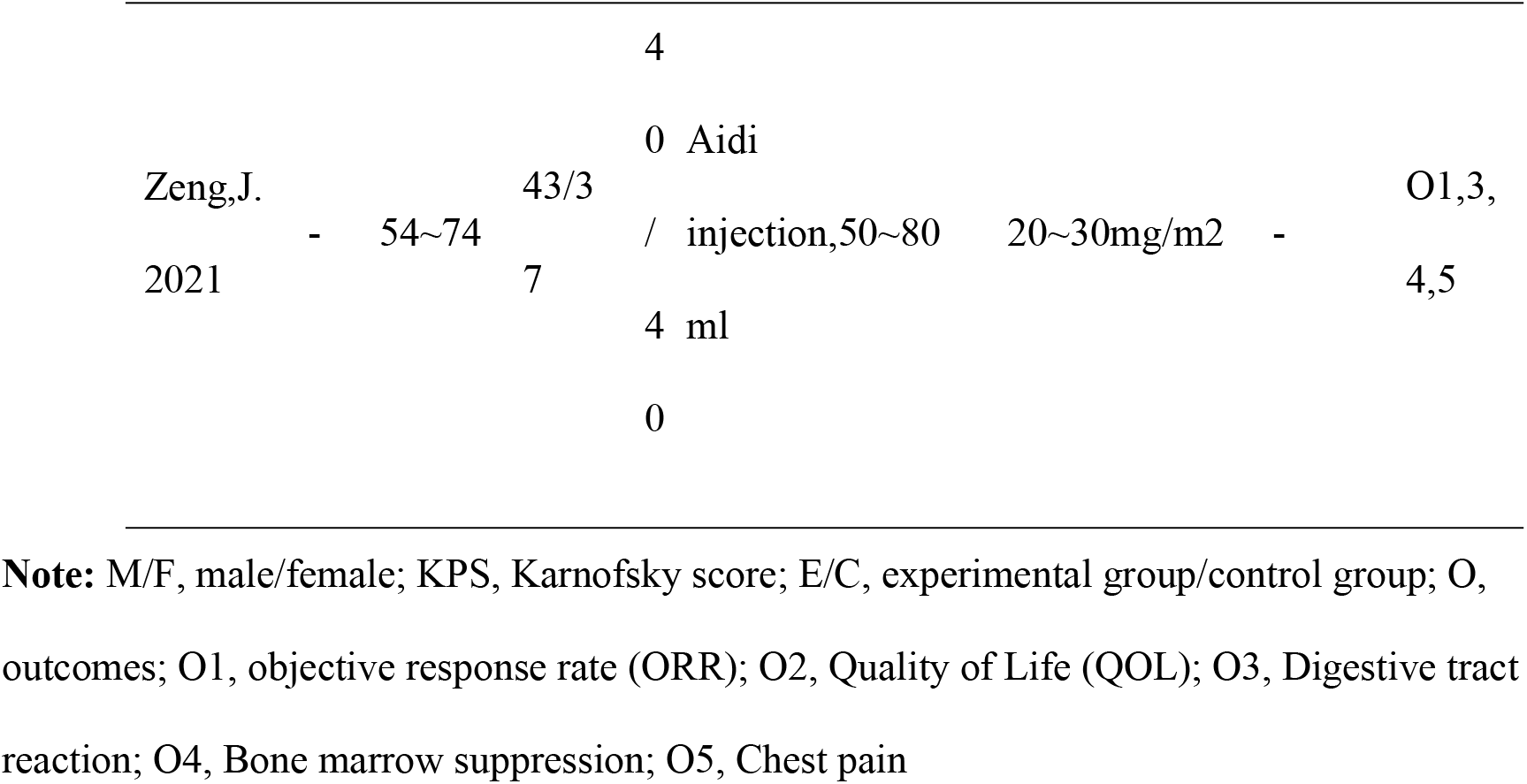
Characteristics of the Included Studies

### 7.3. Study quality and Risk of bias

Among all studies included, 24 used random assignment^[8-10, 15-17, 19-29, 31, 32, 34-36, 38, 39]^, and seven of them reported the way assignments were hidden^[15, 19, 22, 25, 31, 32, 35]^. In terms of blind implementation, none of the studies mentioned it. Two of the included studies may have had incomplete data records, which have no effect on the results of the study ^[20, 40]^, for reasons not explained in the report. In terms of selective reporting of outcomes, all studies reported treatment from two or more aspects, with a low risk of reporting bias. In terms of other biases, 2 treatment methods of the experimental groups were not exclusively described in 2 studies, which may have other biases^[15, 38]^. Figure 2 shows more details. (LocateFigure2)

**Figure 2.**
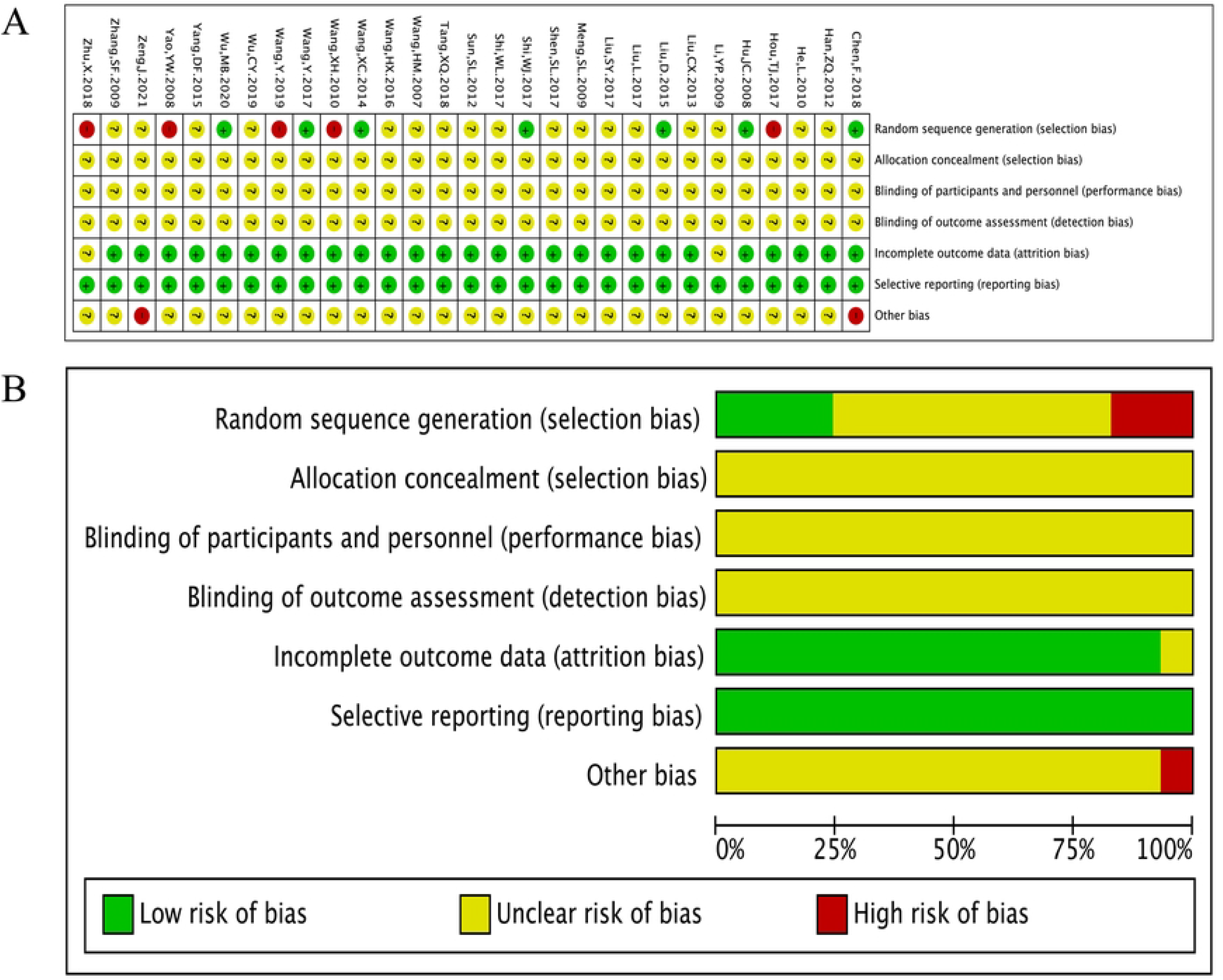
Risk of bias summary and diagram. (A) Risk of bias summary. (B) Risk of bias diagram.

### 7.4. Primary outcomes

#### 7.4.1 Objective Remission Rate (ORR)

Comparing to cisplatin thoracic perfusion, the ORR favored Chinese herbal injection group (RR=1.44, 95% CI: 1.35∼1.53, *P* = 0.000; *I*^*2*^ = 38.4%, *P* = 0.020). (LocateFigure 3)

**Figure 3.**
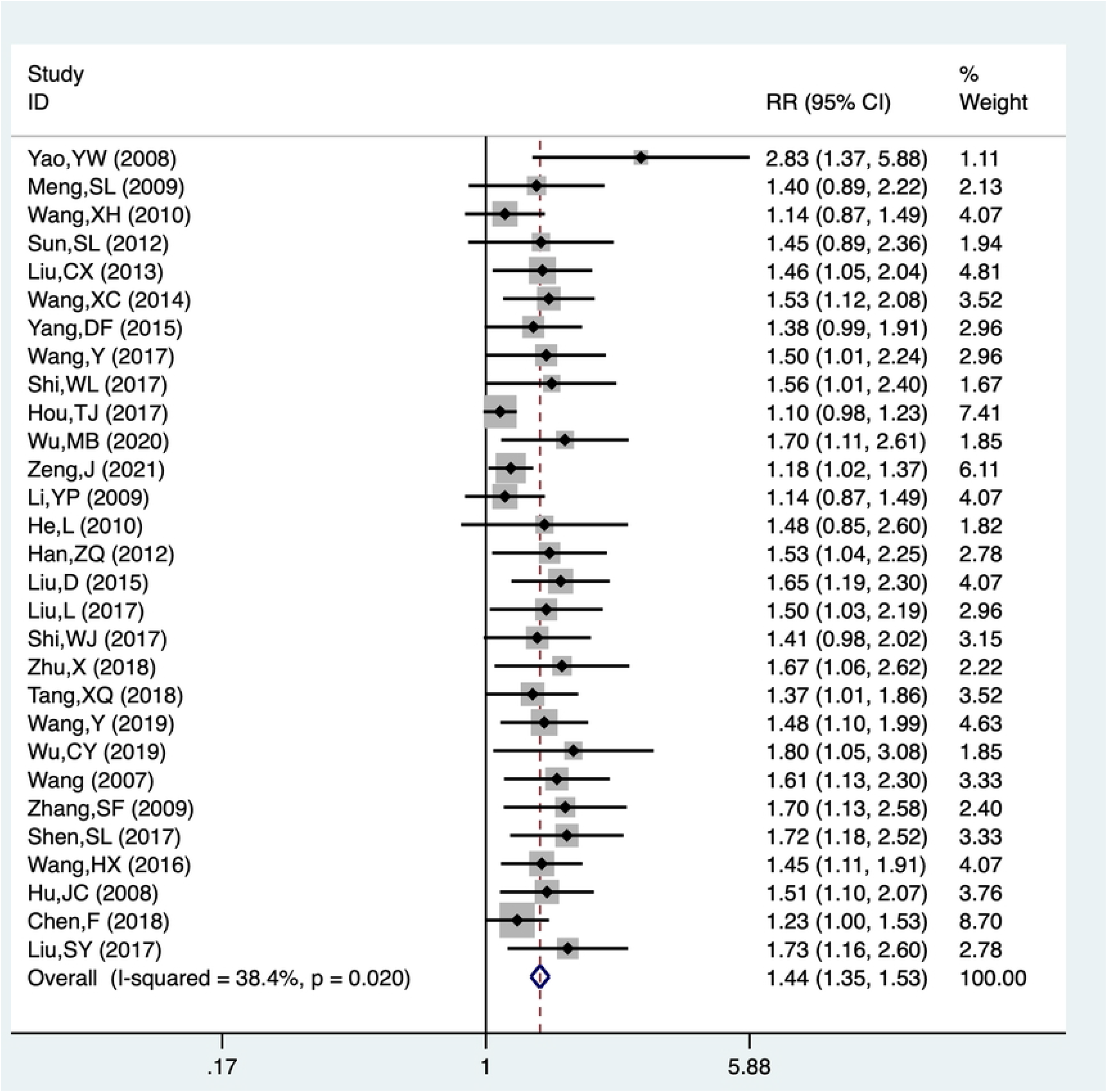
forest plot of ORR

### 7.5. Secondary outcomes

#### 7.5.1 Quality of Life (QOL)

Compared with cisplatin thoracic perfusion alone,Chinese herbal injection thoracic perfusion combined with cisplatin chemotherapy could significantly improve the KPS scores (RR = 1.47, 95%CI: 1.34∼1.61, *P* = 0.0000), with low heterogeneity(*I*^*2*^ = 0%, *P*=0.943), and there was a statistical difference between the two groups. (LocateFigure 4)

**Figure 4.**
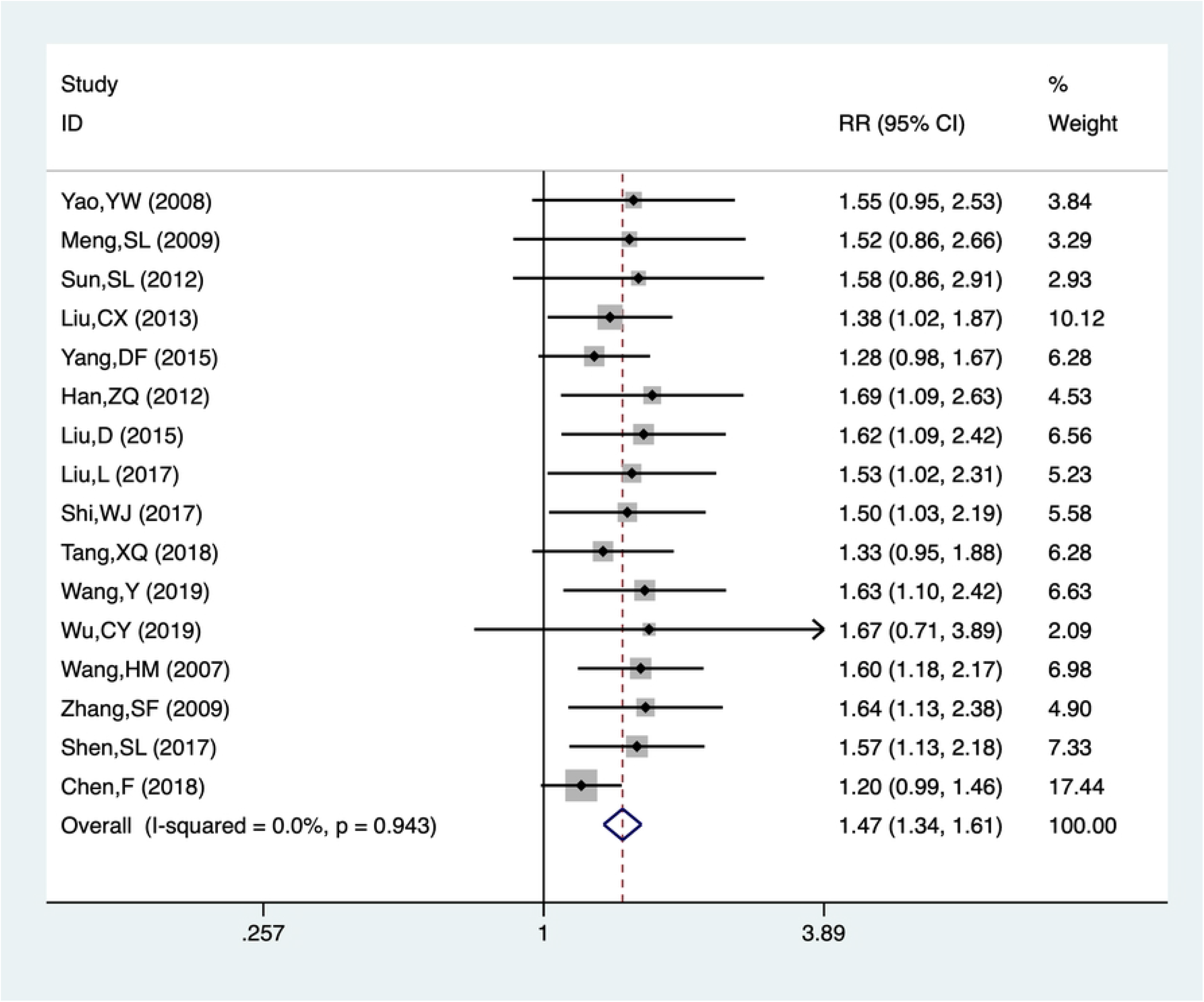
Meta-analysis of QOL

#### 7.5.2 Digestive tract reaction

Comparing to cisplatin thoracic perfusion,Chinese herbal injection thoracic perfusion combined with cisplatin chemotherapy significantly reduced the incidence of digestive tract reactions (RR = 0.56, 95%CI: 0.48∼0.67, *P* = 0.000) with low heterogeneity (*I*^*2*^ = 27.5%, *P* = 0.106). (LocateFigure 5)

**Figure 5.**
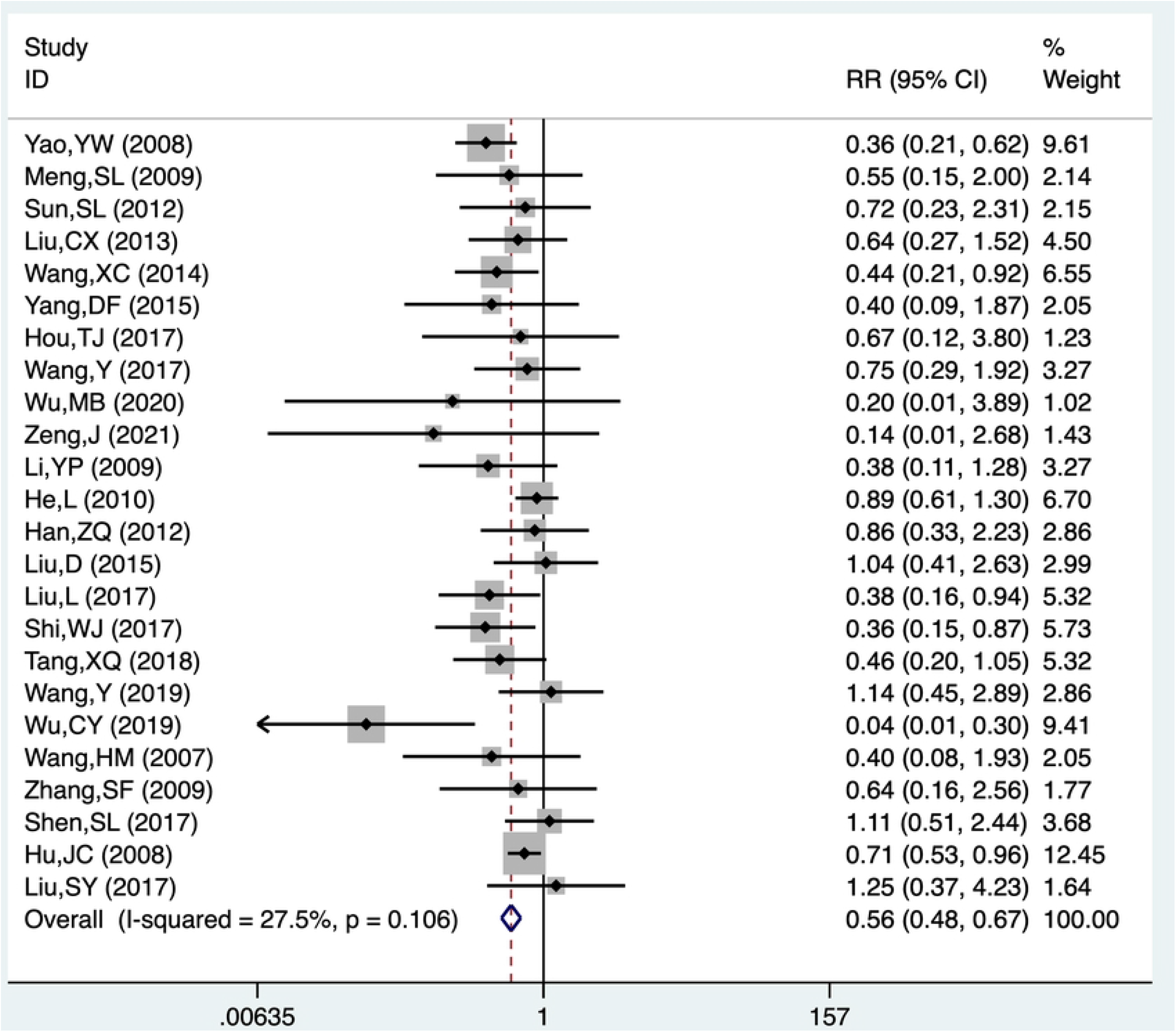
Meta-analysis of digestive tract reaction

#### 7.5.3 Bone marrow suppression

The incidence of bone marrow depressions in experimental group was significantly ower than that in control group (RR = 0.50, 95% CI: 0.43∼0.59, *P* =0.000) with low heterogeneity (*I*^*2*^ = 8.9%, *P* = 0.340). (LocateFigure 6)

**Figure 6.**
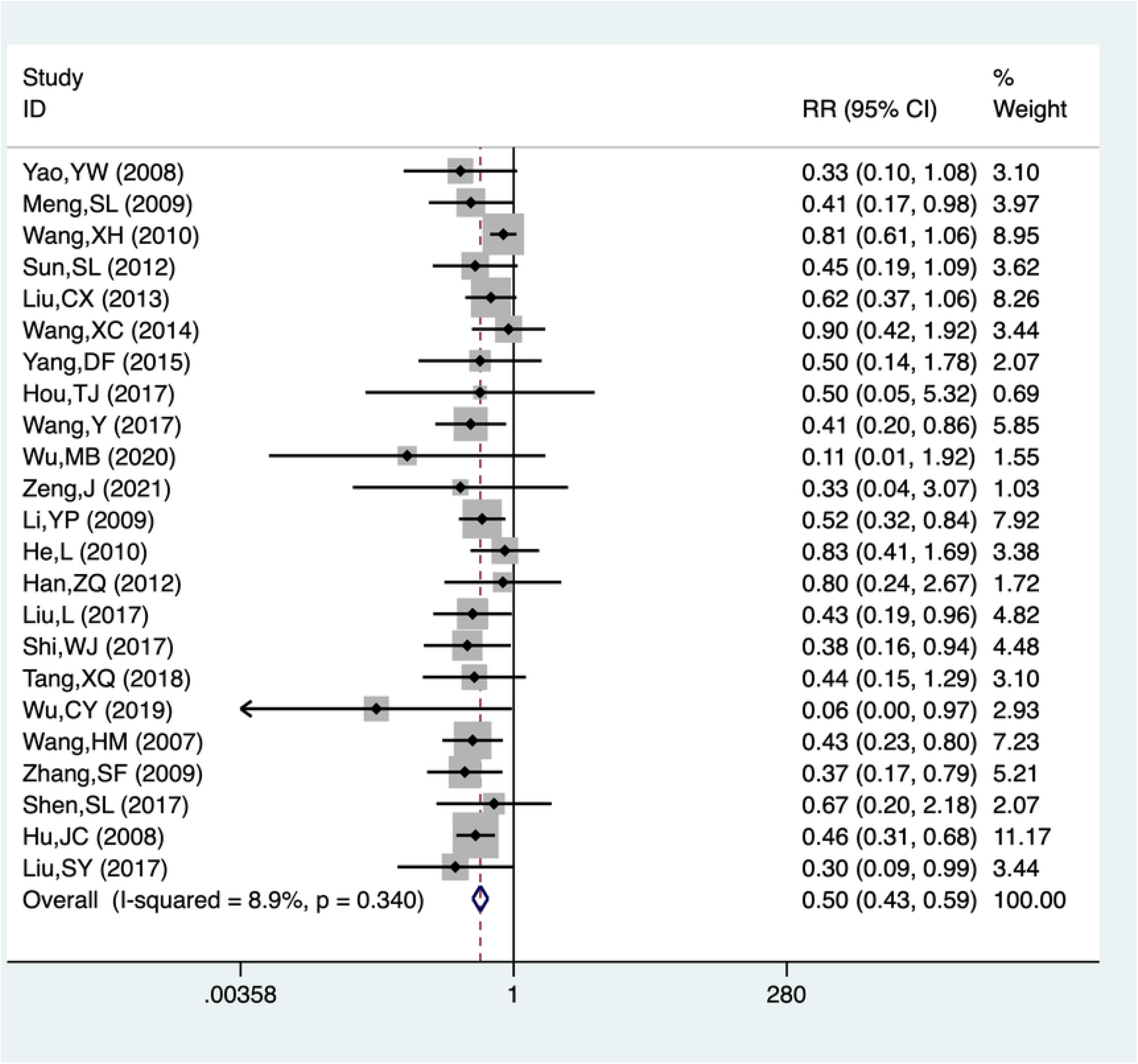
Meta-analysis of bone marrow suppression

#### 7.5.4 Chest pain

The experimental group significantly reduced the incidence of chest pain reactions (RR = 0.65, 95%CI: 0.47∼0. 89, *P* = 0.007), with low heterogeneity (*I*^*2*^ = 0%, *P* = 0.631). (LocateFigure 7)

**Figure 7.**
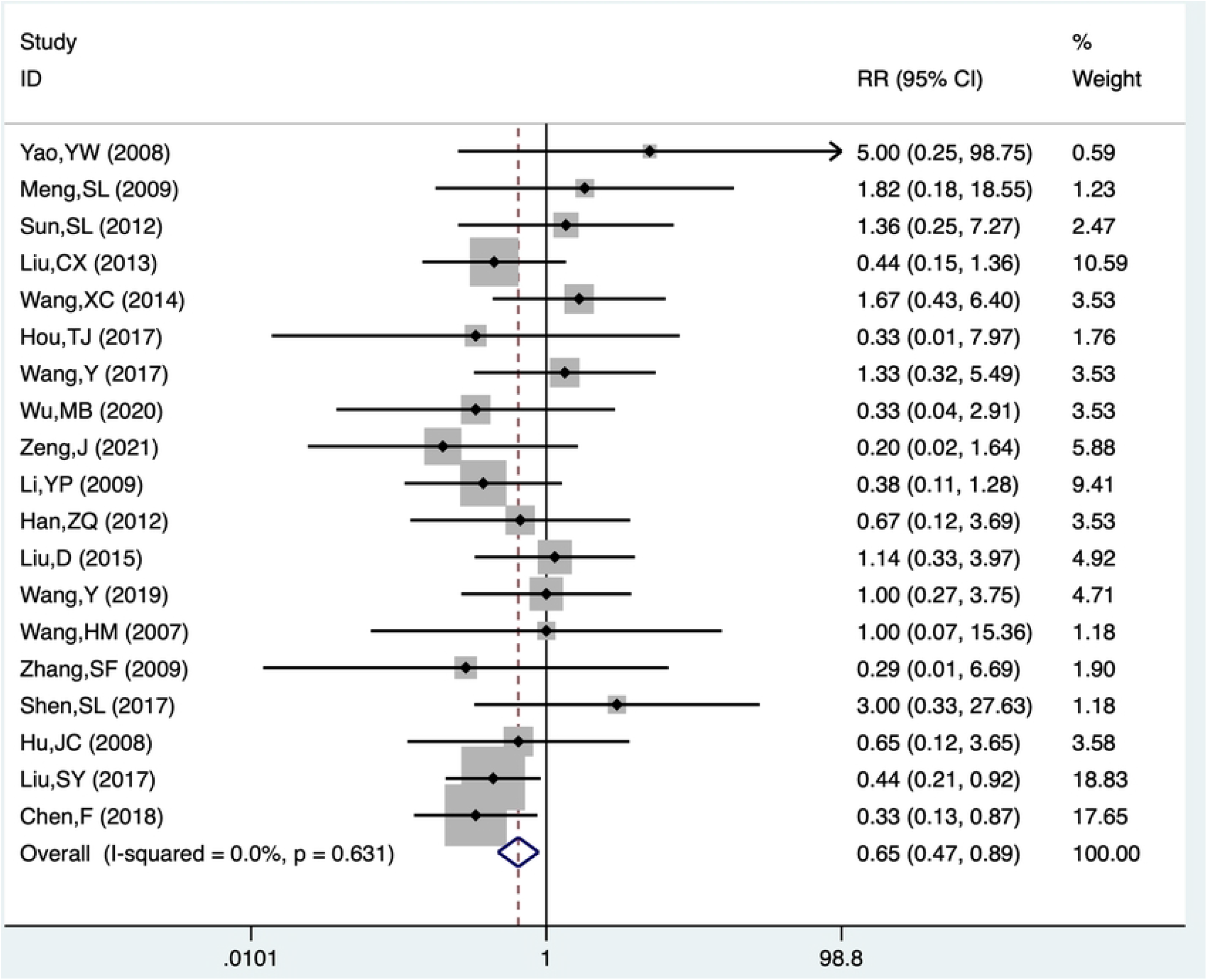
Meta-analysis of Chest pain

### 7.6. Subgroup analysis of QOL and Adverse reactions

Among the 5 secondary outcome measures, subgroup analysis was performed according to the difference of Chinese herbal injections. The results of subgroup analysis are shown in Table 2. Compared with cisplatin thoracic perfusion alone, Aidi injection, Kushen injection, Brucea Javanica oil emulsion combined with cisplatin thoracic perfusion in the treatment of MPE has a better therapeutic effect in QOL, digestive tract reaction, and bone marrow suppression outcome, and the differences are statistically significant, the results of subgroup analysis were consistent with the trend of the overall combined results. But there was no statistically significant difference in the outcome index of chest pain, which was different from the overall combined trend. (Table2)

**Table 2.** results of subgroup analysis

| Outcome | Pooled<br>RR(95%CI) | Z | P | Heterogeneity |  |
| --- | --- | --- | --- | --- | --- |
| | | | | $I^2(\%)$ | $P_h$ |
| Quality of<br>Life (QOL) | 1.52(1.37,1.69) | 8.05 | 0.000 | 0% | 0.998 |
| Digestive<br>tract<br>reaction | 0.53(0.43,0.64) | 6.37 | 0.000 | 27.9% | 0.111 |
| Bone marrow suppression | 0.52(0.44,0.61) | 7.60 | 0.000 | 8.6% | 0.346 |
| Chest pain | 0.82(0.54,1.25) | 0.93 | 0.35 | 0% | 0.729 |

### 7.7. Moderator effects

To investigate the sources of potential heterogeneity, we conducted subgroup analysis with moderator variables include injection categories, According to the results of subgroup analysis in Figure 8, it can be concluded that Aidi injection combined with cisplatin thoracic perfusion (RR=1.37, 95%CI: 1.25∼1.50, *P* = 0.000, *I*^*2*^ =58.3%, *P*=0.006), Kushen injection combined with cisplatin thoracic perfusion (RR = 1.48, 95%CI: 1.32∼1.66, *P* = 0.000, *I*^*2*^ = 0%, *P*=0.813). Brucea Javanica oil emulsion combined with cisplatin thoracic perfusion was infused into the chest (RR = 1.68, 95%CI: 1.34∼2.09, *P* = 0.000, *I*^*2*^ = 0%, *P*=0.964). Compared with cisplatin thoracic perfusion alone, all of them have more effective rate in the treatment of MPE, and the difference is statistically significant. Therefore, the results of the subgroup analysis on ORR, the primary outcome measure, were trend consistent with the overall pooled results (RR = 1.45, 95%CI: 1.35∼1.55, *P* = 0.000, *I*^*2*^ = 45.2%, *P*=0.008). (LocateFigure8)

**Figure 8.**
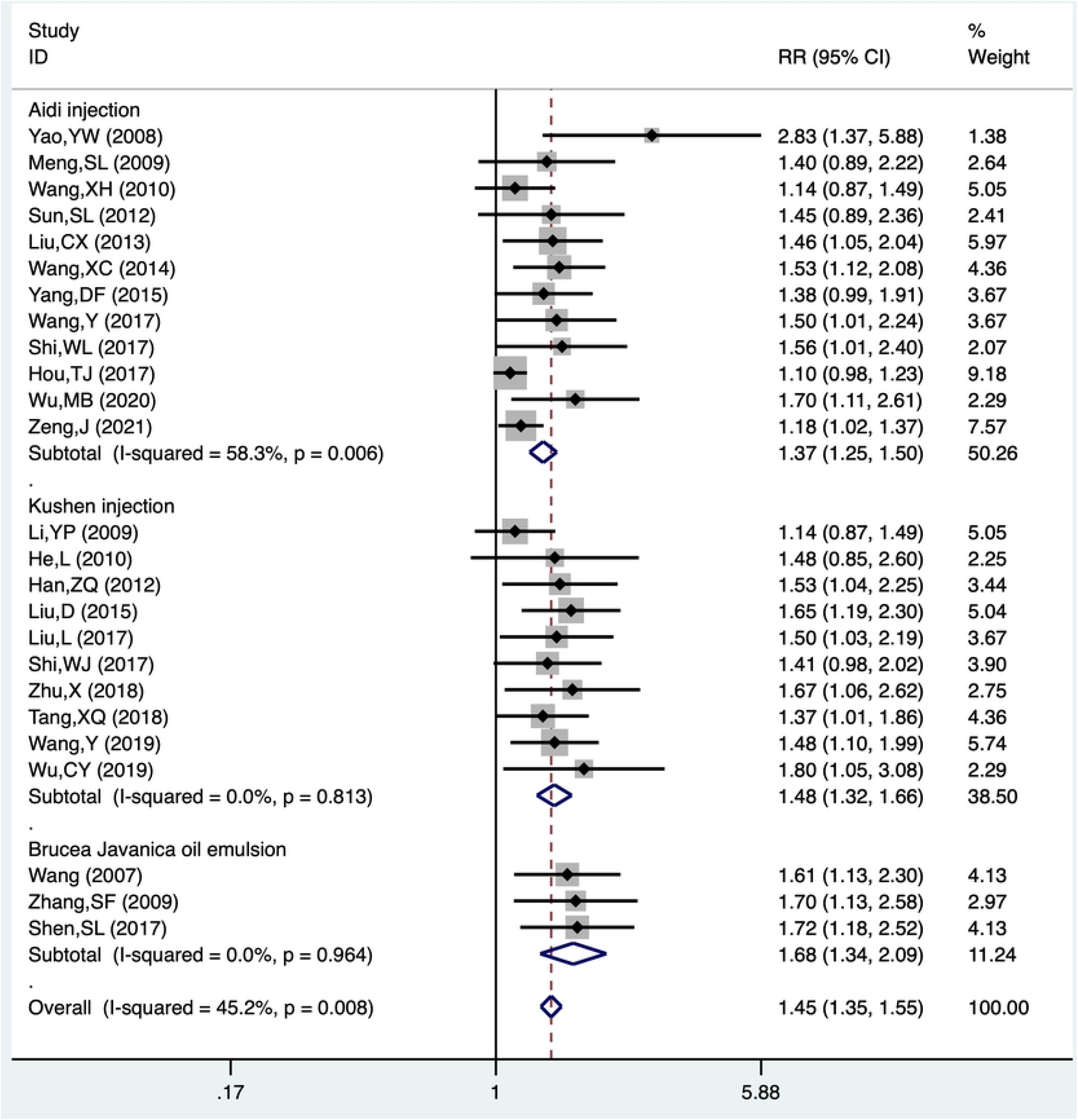
Subgroup analysis of ORR

### 7.8. Sensitivity analysis

In this study, sensitivity analysis was conducted leave-one-out method, revealing that Hou TJ^[18]^ had a rather great impact on heterogeneity. After the elimination, *I*^*2*^ decreased from 38.4% to 0%, but had little effect on the final results (RR=1.44, 95% CI: 1.35∼1.53, P = 0.000 vs RR:1.47, 95%CI: 1.37∼1.56, P = 0.0000). There was a certain heterogeneity in subgroup analysis of ORR (*I*^2^ = 45.2%). After deleting the study of Hou TJ^[18]^, *I*^*2*^ decreased from 45.2% to 0%, but had little effect on the final results (RR = 1.42, 95%CI: 1.30∼1.55, *P* = 0.000 vs RR:1.40, 95%CI: 1.31∼1.50, P = 0.0000). (Figure 9A-B). Compared with other research, Hou TJ had a relatively high response rate, so as to tilting the heterogeneity much heavier. For second outcome: Quality of life (QOL), digestive tract reaction, bone marrow suppression and chest pain, outcome indicators found that the heterogeneity among studies was small (I^2^ <30%), and the inclusion of one study by one had little impact on the results, so these results were stable (LocateFigure 9).

**Figure 9.**
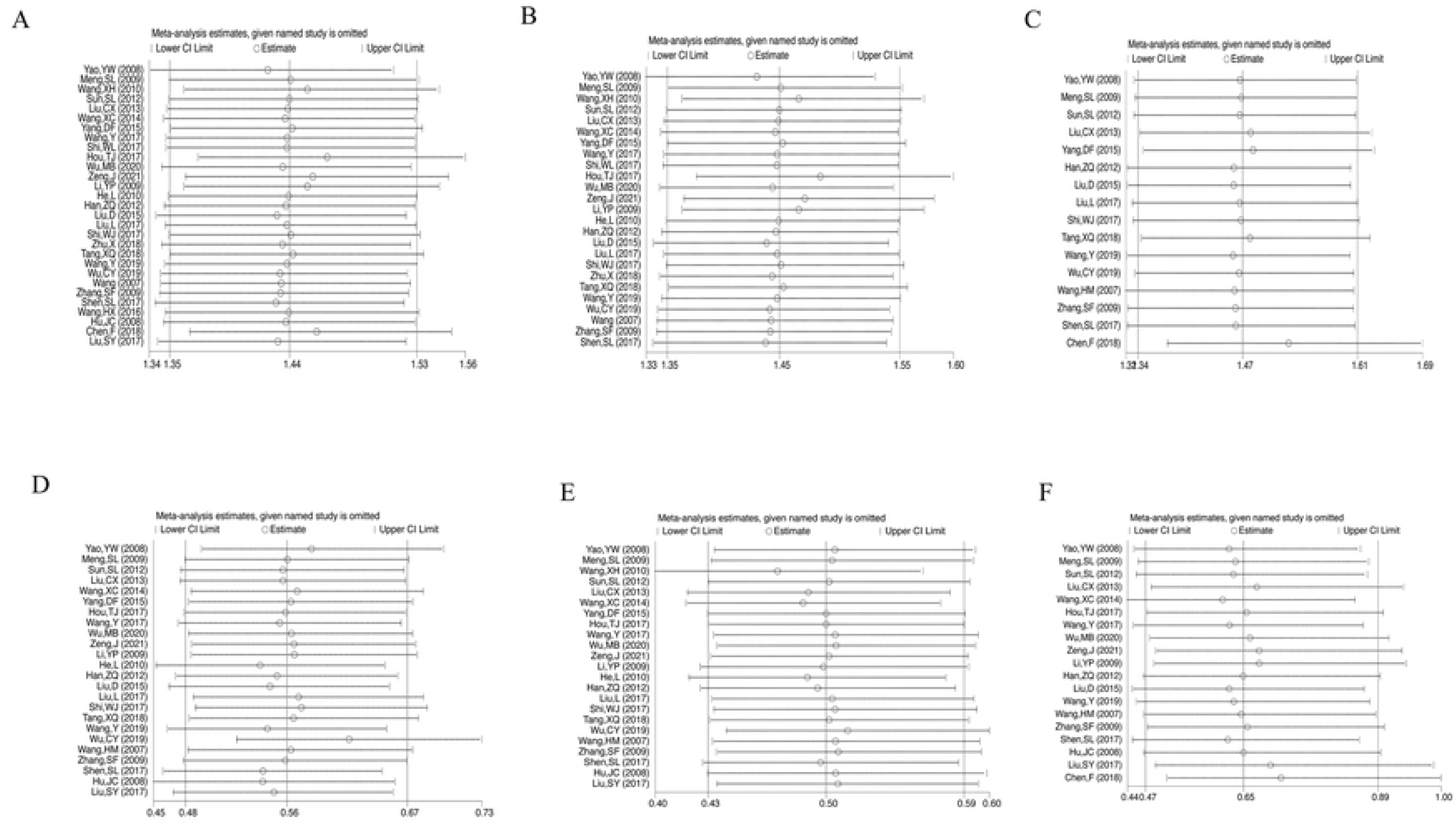
(A) sensitivity analysis of ORR; (B) sensitivity analysis of Subgroup analysis of ORR; (C) sensitivity analysis of QOL; (D) sensitivity analysis of digestive tract reaction; (E) sensitivity analysis of bone marrow suppression; (F) sensitivity analysis of chest pain

### 7.9. Publication bias

Funnel plots were drawn according to the primary outcome measure ORR, we noticed that the included study was distributed asymmetrically on both sides of the funnel chart (Figures 10A–C), indicating that there was significant publication bias in the meta-analysis.The Begg (p=0.003) and Egger (p=0.000) test was used to test the publication bias of 29 studies included in this meta-analysis, and the results showed that this study had obvious publication bias (P<0.05). The 29 articles included in this study were analyzed by the trim-fill method, and the results showed that after the inclusion of 14 virtual studies, Meta-analysis was conducted again, using the fixed effect model, and the combined results of effect indicators RR=1.229, 95%CI :1.171∼1.289, although some small sample studies were added to correct the publication bias, which did not have a significant impact on the results. (LocateFigure10)

**Figure 10.**
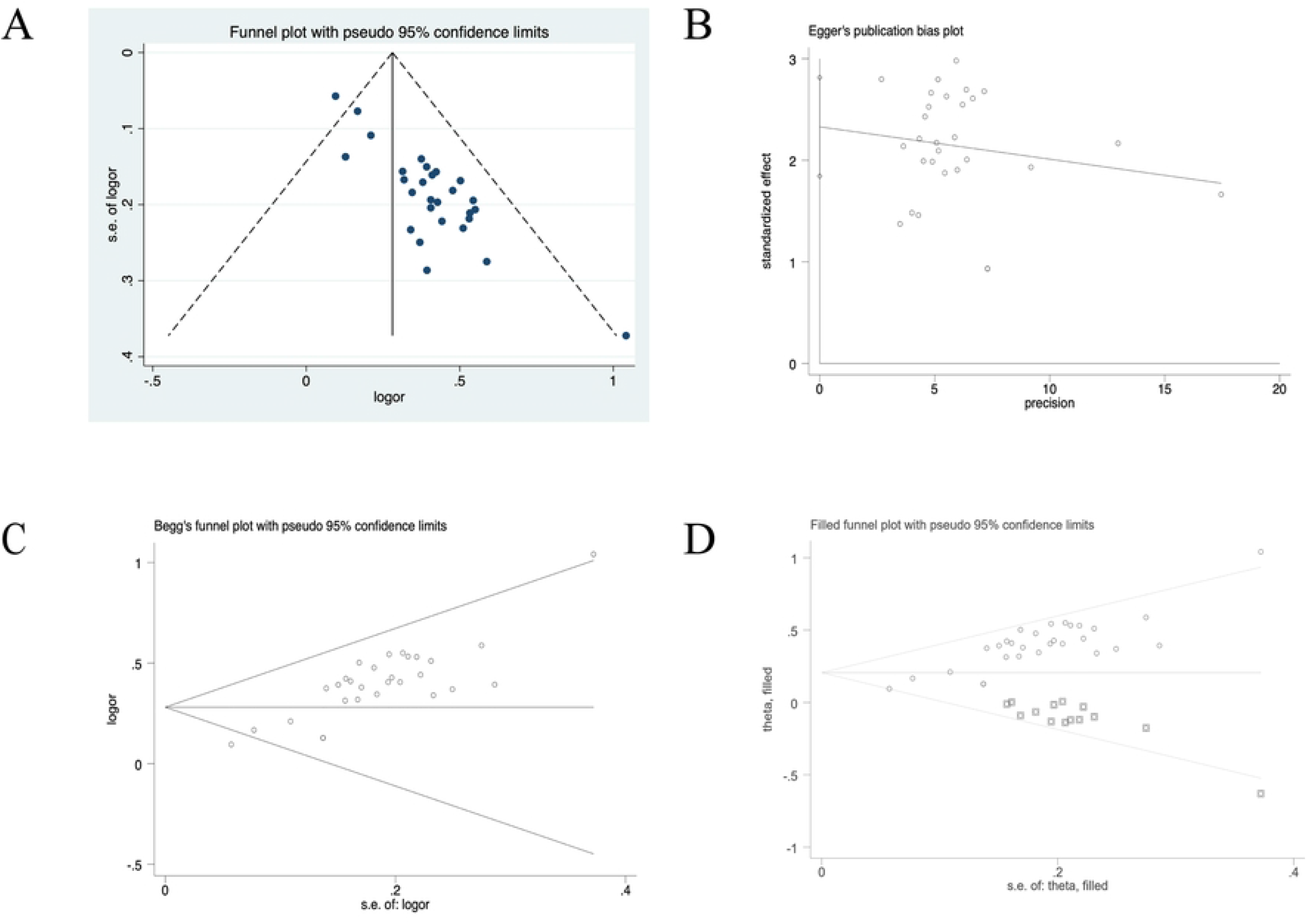
(A) Funnel plot of ORR; (B) Egger test of ORR; (C) Begg test of ORR; (D) trim-fill method of ORR

## 8. Discussion

The aim of this meta-analysis is to evaluate the efficacy and safety of Chinese herbal injection thoracic perfusion combined with cisplatin chemotherapy in the treatment of lung cancer MPE. Compared with the published meta-analysis^[42]^, our study is the first to control the administration mode of traditional Chinese herbal injection as thoracic perfusion, using thoracic perfusion of cisplatin as the control group, which ensures the consistency of intervention measures and can reduce clinical heterogeneity to a certain extent. Our results demonstrated that Chinese herbal injection thoracic perfusion combined with cisplatin chemotherapy can improve the ORR of lung cancer patients with MPE (RR=1.44, 95% CI: 1.35∼1.53, *P* = 0.000). In addition, we also performed subgroup analysis according to different Chinese herbal injection. The results showed that compared with cisplatin thoracic perfusion alone, Aidi injection, Kushen injection and Brucea Javanica oil emulsion could improve the ORR of lung cancer patients with MPE.

As we all know, lung cancer patients with MPE will have symptoms of chest pain due to poor water circulation. Chemotherapy drugs will inhibit the hematopoietic function of bone marrow, and also affect the digestive function of patients, with vomiting, diarrhea and other digestive tract reactions, further affecting the quality of life of patients. In terms of quality of life, our results showed that Chinese herbal injection could effectively improve the quality of life of patients with lung cancer MPE, and the results of subgroup analysis were consistent with the total results. In addition, Chinese herbal injection thoracic perfusion combined with cisplatin chemotherapy can effectively improve the digestive tract reaction and bone marrow suppression of patients, and the results are statistically significant and consistent with the results of subgroup analysis. Chinese herbal injection thoracic perfusion combined with cisplatin chemotherapy can alleviate chest pain in patients, but the results of subgroup analysis showed no statistical significance, which may be related to the small sample size included in the study. We should consider focusing on this adverse reaction in the future.

Although funnel plots for the 29 studys showed that there was significant publication bias in the meta-analysis, there is no significant difference between the results of trim-fill method and the original data, which is favorable to our existing results

Our study also have certain limitations, which increase the risk of bias in the outcome indicators of this study. First of all, although this study adopted a relatively extensive search strategy, some conference papers, supplements and grey literature may not be fully obtained, so potential publication bias cannot be excluded. The funnel plot and Begg and Egger test results also reflect some potential publication bias. Among the 29 included studies, only 7 studies described the method of randomization, while the remaining studies did not explain the method of randomization and allocation concealment, and no study mentioned the blinding method, which may lead to potential performance bias and selection bias. Secondly, the sample size of all the included studies was generally small, and the efficacy of the subjects was not tracked. The intervention measures, intervention time and drug administration plan of all the studies were not completely unified, and the forms and effectiveness of the intervention mechanisms could not be clarified. It may increase the imprecision of the outcome indicators and make the evidence strength of the outcome indicators low. Finally, important clinical survival outcomes such as overall survival, progression-free survival, and 5-year survival were not fully reported in the included studies. Therefore, high quality and large sample clinical studies are needed in the future to further evaluate the efficacy and safety of Chinese herbal injection thoracic perfusion combined with cisplatin chemotherapy in the treatment of lung cancer MPE, and provide more evidence to support our findings.

## 9. Conclusion

Our study demonstrated the potencies of Chinese herbal injection thoracic perfusion combined with cisplatin chemotherapy to enhance the efficacy and safety for lung cancer MPE patients, improve quality of life and decrease adverse reactions. Furthermore, the long-term efficacy of Chinese herbal injection in Lung cancer MPE treatment still needs to be verified in future well-designed clinical trials that adhere to CONSORT guidelines. More efforts are needed to promote the application of Chinese herbal injection in the clinic.

## Data Availability

All relevant data are within the manuscript and its Supporting Information files.

## Acknowledgments

We sincerely thank all support from all authors.

## Data Availability

The datasets generated during and/or analysed during the current study are available from the corresponding author on reasonable request

